# Deep Learning for Individual-Level Classification of Schizophrenia Versus Healthy Controls from Trial-Level Auditory Oddball ERP Waveforms

**DOI:** 10.64898/2026.07.24.26358816

**Authors:** Yi-Han Sheu, Yi-Ting Lin, Kristina M. Holton, Chih-Min Liu, Yi-Ling Chien, Chen-Chung Liu, Mei-Hua Hall, Hai-Gwo Hwu, Ming H. Hsieh

## Abstract

Machine learning approaches may support individual-level classification in psychiatry, but many EEG-based schizophrenia studies have relied on small samples or conventional summary features. We evaluated whether trial-level auditory oddball event-related potential (ERP) waveforms could support schizophrenia versus healthy-control classification using deep learning. The study included 258 patients with schizophrenia and 142 healthy controls. EEG recordings from an auditory duration oddball paradigm were segmented into −100 to 500 ms epochs, and trial-level mismatch waveforms were generated by subtracting each participant’s mean standard response from accepted deviant trials. Models were trained using a fixed participant-level training, validation, and test split, with demographic residualization fit only in the training set. Five deep learning architectures were trained on full residualized ERP waveforms and compared with classical machine learning models trained on 18 conventional ERP summary features. Deep learning models achieved higher test set discrimination than classical feature-based models, with AUROC values ranging from 0.797 to 0.857 versus 0.705 to 0.720. Benchmark analyses suggested that performance depended on the combination of waveform-level input and deep learning architecture. These findings support trial-level auditory oddball ERP waveforms as promising classification inputs and candidate electrophysiological biomarkers of schizophrenia-related neural information processing.

## Introduction

Machine learning (ML) methods are increasingly applied in medicine to support prediction and classification from complex, high-dimensional data^1,2^. In psychiatry, these approaches have been used to develop computational models for tasks including risk stratification, treatment-response prediction, and diagnostic prediction or patient classification^3–5^. Among ML approaches, deep learning (DL), which uses multi-layer neural network architectures to learn representations directly from data, has shown particular promise in domains involving high-dimensional or minimally processed inputs. Despite growing interest, the application of these methods in psychiatry remains an emerging area of research^6^.

Schizophrenia is a severe and heterogeneous psychiatric disorder associated with abnormalities spanning genetic liability, neurochemical signaling, information processing, and behavior^7^. Within precision psychiatry, there is growing interest in models that support inference at the level of the individual, including single-subject prediction and classification^8,9^. To date, ML and DL approaches in schizophrenia research have been applied predominantly to genomic data, electronic health records, and structural or functional magnetic resonance imaging (MRI), whereas electrophysiological biomarkers such as event-related potentials (ERPs) have received comparatively less attention^10–12^. This gap is notable because, although MRI-based approaches have shown promise, they remain constrained by cost, accessibility, and limited temporal resolution for resolving the rapid neural dynamics relevant to schizophrenia^6,13^. In contrast, auditory oddball ERPs provide a low-cost, temporally precise, and experimentally controlled measure of neural information processing. Resting-state EEG^14^ is discriminative but is less often directly tied to a specific cognitive operation than stimulus-locked ERPs, which index defined neurocognitive processes. ERPs can therefore serve not only as inputs for classification models but also as operationalizable biomarkers of disease-relevant brain function.

Mismatch negativity (MMN) and P3a ERPs elicited using a passive oddball paradigm represent promising electrophysiological markers of schizophrenia because they reflect stimulus-locked neural responses associated with sensory prediction, deviance detection, and attentional orienting. MMN is a negative deflection occurring approximately 100–250 ms after auditory deviance and is associated with echoic memory and automatic deviance detection, whereas P3a is associated with involuntary attentional orienting^15,16^. Duration MMN is regarded as a strong candidate endophenotype for bridging the gene-to-phenotype gap for schizophrenia^16,17^. MMN reductions constitute one of the most robust neurophysiological findings in schizophrenia: meta-analyses have reported large standardized effect sizes (0.95–0.99), with deficits generally greater in chronic than in first-episode schizophrenia^18,19^. MMN deficits are highly heritable, evident in attenuated responses in unaffected first-degree relatives Duration MMN is heritable and is attenuated in unaffected first-degree relatives^20,21^, and associated with subsequent transition to psychosis in high-risk individuals^19,22^. Mechanistically, MMN deficits have been linked to N-methyl-D-aspartate receptor (NMDAR) hypofunction and excitation–inhibition imbalance^23,24^. Reduced P3a amplitude is also a robust finding in schizophrenia^25^, including in first-episode psychosis^26^. A key advantage of MMN and P3a is that they can be elicited from a passive oddball paradigm, minimizing reliance on active task performance and reducing confounding related to impaired motivation or cooperation^9,16^.

Although abnormalities in these ERPs are well documented at the group level, their utility for individual-level classification remains limited. One potential reason is that conventional ERP analyses are typically based on summary features extracted from averaged waveforms, such as peak amplitude and latency, which may obscure informative temporal structure and trial-to-trial variability^27^. Preserving this information may be important for developing models that distinguish patients with schizophrenia from healthy controls at the individual level.

Deep learning models offer a promising approach for overcoming this limitation by learning complex, non-linear structure from raw or minimally processed time-series data^13,14^. Traditional machine learning methods, by contrast, often struggle when applied directly to full waveforms because of their high dimensionality, noise, and temporal variability, and therefore typically rely on hand-engineered features such as averaged peak amplitudes or latencies. Deep learning architectures instead automatically learn hierarchical representations directly from high-dimensional inputs^12,28^, potentially capturing subtle morphological and temporal characteristics that are attenuated or lost in conventional summary-based analyses. However, few studies have systematically evaluated whether deep learning models trained on full trial-level auditory oddball ERP waveforms can support individual-level schizophrenia classification, or whether classification performance depends primarily on model architecture, input representation, or their combination. Despite promising results from prior EEG-based schizophrenia classifiers, studies have relied on relatively small cohorts or repeatedly used public datasets^14^, limiting confidence in the generalizability of reported performance estimates.

Using a comparatively large case-control sample of 258 patients with schizophrenia and 142 healthy controls, the present study examined whether auditory oddball ERPs can support individual-level schizophrenia-versus-healthy-control classification. We first evaluated multiple deep learning architectures, including multi-layer perceptrons, convolutional neural networks, recurrent neural networks, autoencoders, and transformers, trained directly on full residualized trial-level mismatch waveforms to preserve waveform-level information beyond conventional averaging. We then compared their performance with that of traditional machine learning models trained on features derived from intra-subject averaged ERPs. As complementary benchmarks, we evaluated deep learning models trained on the same 18 ERP summary features and traditional machine learning models trained on full residualized trial-level mismatch waveforms. This design allowed us to assess whether classification performance was driven primarily by model architecture, input representation, or their combination.

## Methods

### Study Participants

The study included 258 patients with schizophrenia and 142 healthy controls recruited from National Taiwan University Hospital. All participants provided written informed consent, and the study protocol was approved by the institutional review board of National Taiwan University Hospital (approval numbers 200812098R, 201312111RIN, and 201903116RIN). Clinical and demographic assessments included age, sex, illness duration, age at illness onset, smoking status, and symptom severity as assessed by the Positive and Negative Syndrome Scale (PANSS). Differences in demographic characteristics between diagnostic groups were quantified using the chi-square test for categorical variables and Cohen’s d for continuous variables, where applicable.

### Recording environment and auditory oddball paradigm

32-lead EEG was recorded with electrode placement following the International 10–20 system. Electrodes placed at the tip of the nose served as the reference, and Fpz as the ground. Four additional electrodes were positioned above and below the left eye and at the outer canthi of both eyes to monitor blinks and eye movements, in addition to an electrocardiography channel and a reference channel. All electrode impedances were maintained below 5 kΩ. Signals were digitized at 1 kHz with system acquisition filter settings of 0.5 to 100 Hz, without a 60-Hz notch filter.

The standard passive auditory oddball paradigm^29,30^ was employed in accordance with protocols previously established in our laboratory and used in prior studies^31–37^. Auditory stimuli consisted of 1000 Hz tones at 80 dB with a 1-ms rise-fall time, presented at a fixed 500-ms interstimulus interval. We applied a duration-deviant auditory oddball paradigm, in which standard tones (probability = 0.90; 50-ms duration) and deviant tones (probability = 0.10; 100-ms duration) were presented in a pseudorandom sequence, ensuring at least two standards preceded each deviant. Subjects were instructed to relax with their eyes open and to focus on watching a silent cartoon on a video monitor during the passive auditory oddball paradigm. Auditory stimuli were generated and presented to subjects binaurally via foam insert earphones during the silent cartoon. Online recording continued until a minimum of 225 artifact-free deviant trials were obtained, with the entire session lasting approximately 20 to 30 minutes (a minimum of 2250 total trials).

### ERP processing

Offline EEG data processing was conducted using automated routines by personnel blinded to subject clinical group. Ocular artifact correction was applied first. Continuous EEG recordings were segmented into epochs from −100 to 500 ms relative to stimulus onset, thereby capturing the MMN and P3a time windows. Epoch files were subsequently low pass filtered at 40 Hz, linearly detrended, and baseline corrected using the pre stimulus interval. The exported data contained a total of 36 channel traces, including 32 EEG channels, two electrooculography channels (derived by the differences in the horizontal and vertical ocular channels), the electrocardiography and reference channels. Because these non-scalp traces were present in the exported recordings and may capture reference-related or common-mode variation, they were retained as auxiliary recording channels and provided to the models together with the scalp EEG channels rather than excluded a priori.

### Calculating grand-average ERP measures

Traditional grand-average ERPs were calculated and compared between the case and control groups using Student’s t-tests. Separate average waveforms were first computed for the standard and deviant stimuli. The ERP waveform elicited by the standard stimuli was then subtracted from that elicited by the deviant stimuli to derive the MMN/P3a difference waveform. MMN and P3a were quantified based on their peak amplitudes and latencies within the 90–250 ms and 210–350 ms time windows, respectively. Based on prior literature identifying Fz as the site of maximal MMN response, this electrode was selected for the between-group comparison of MMN.^30^

### Epoch-level data for waveform-based analysis

To prepare the data for waveform-based analysis, a trial-level mismatch waveform was derived for each accepted deviant trial by subtracting the participant’s mean standard-trial waveform from the individual deviant-trial waveform. Averaging the standard-trial waveforms provided a stable, participant-specific baseline for calculating the time-locked ERP difference between deviant and standard responses, while retaining the within-subject variability and trial-level information present in the individual deviant trials. Each resulting trial was represented as a 36 × 601 channel-by-time matrix for model input, preserving spatial and temporal information across the epoch. The number of matrices per participant therefore corresponded to that participant’s number of accepted deviant trials.

### Training, validation, and test split

Participants were partitioned once into non-overlapping training (n = 250), validation (n = 50), and test (n = 100) sets. The split was used identically by every model in this study. All hyperparameter selection and decision-threshold locking occurred on the training and validation subjects only; the 100 test participants were excluded during model development and were only scored after all training, tuning, and threshold selection were complete.

### Feature representations

Two parallel feature representations were used, each matched to the natural input structure of the corresponding model family. The first representation consisted of full residualized waveforms (see below), with each trial represented as a 36 × 601 channel-by-time matrix. When flattened, this representation yielded 21,636 features per trial. This waveform-level input was used by all five deep learning models and by the two tree-based classical machine learning models that were computationally tractable at this dimensionality.

The second representation consisted of residualized ERP summary features, with each participant represented by an 18-dimensional feature vector. These features included 12 peak-based measures: MMN and P3a mean peak latencies and amplitudes at Fz, FCz, and Cz. They also included four mean-area amplitude features computed over the canonical MMN and P3a time windows at Fz and FCz, as well as two frontocentral average MMN amplitude features, Fz-FAMMN and FCz-FAMMN.

### Demographic residualization

To reduce the risk that classifiers would exploit demographic differences between patients with schizophrenia and healthy controls rather than illness-specific neurophysiology, all EEG-derived inputs were residualized with respect to age, sex/gender, years of education, and smoking burden before model training. Residualization was performed at the participant level for both input representations. For trial-level waveforms, regression coefficients were estimated from each training participant’s mean waveform, and the resulting demographic correction was applied to all trials from that participant. For the 18 ERP summary features, a separate regression was fitted for each feature using one observation per participant. In both cases, coefficients were estimated using only the 250 training participants and applied unchanged to validation and test participants. Full residualization procedures, imputation details, and diagnostics are provided in the Supplementary Methods A.

### Classification Models

#### Deep learning models on full residualized waveforms

Five deep learning architectures were trained on trial-level residualized EEG waveforms: a transformer, convolutional autoencoder with classification head, convolutional neural network, recurrent neural network, and multi-layer perceptron. These models were selected to represent complementary approaches to waveform classification, including attention-based sequence modeling, convolutional feature learning, recurrent temporal modeling, reconstruction-guided representation learning, and fully connected classification of flattened waveform inputs. Hyperparameters were selected with Optuna using validation-set participant-level AUROC, and early stopping was based on validation performance. The best validation checkpoint for each architecture was retained for held-out test-set evaluation without retraining on the combined training and validation data. Full architecture specifications, tuning parameters, and optimization settings are provided in the Supplementary Methods B and Supplementary Table 1.

#### Classical machine learning models on 18 residualized ERP summary features

Four classical machine learning classifiers were trained on the 18 residualized ERP summary features: penalized logistic regression (elastic net), support vector classification with linear and radial basis kernels, random forest, and gradient boosting. Hyperparameters were selected by grid search with 5-fold stratified cross-validation within the training split, using AUROC as the model selection criterion. Linear and support vector models included feature standardization within the cross-validation pipeline, whereas tree-based models were trained on unstandardized features. Full model specifications and hyperparameter grids are provided in the Supplementary Methods C and Supplementary Table 2.

#### Multi-layer perceptron probe on 18 residualized ERP summary features

To isolate the contribution of model architecture from input representation, we trained an additional MLP on the 18 participant-level ERP summary features using the same training, validation, and test partitions. The MLP was selected from the five deep learning architectures because its relatively simple fully-connected structure imposes fewer assumptions about temporal locality or sequential dependence than convolutional, recurrent, or attention-based models, making it a suitable neural-network comparator for low-dimensional summary features. We evaluated two variants: a “scaled” MLP with a search space tailored to the 18-feature input and a “large” MLP with a search space matched to the waveform-based MLP. This analysis tested whether neural-network architecture alone improved performance when models were restricted to conventional ERP summary inputs rather than full waveform epochs.

Hyperparameters were selected with Optuna on the validation split, and the best validation checkpoint was evaluated on the held-out test set. Full search spaces and implementation details are provided in Supplementary Methods D and Supplementary Table 3.

#### Classical machine learning models on full residualized waveforms

As a complementary benchmark, we trained classical tree-based ensemble models on flattened residualized waveform inputs using the same training, validation, and test partitions. This analysis tested whether access to full epoch-level waveform information alone was sufficient to reproduce the performance of deep learning models. Hyperparameters were selected using participant-grouped cross-validation within the training split, and participant-level predictions were obtained using the same trial-aggregation procedure applied to the waveform-based deep learning models. Penalized logistic regression and linear support vector models were also attempted as linear full-waveform comparators but did not converge or complete fitting within the available compute budget, and therefore were not included in the final quantitative comparisons. Full hyperparameter grids, implementation details, and computational feasibility analyses are provided in the Supplementary Methods E and Supplementary Table 4.

### Threshold selection, Evaluation metrics, and Uncertainty quantification

Decision thresholds were selected on the validation split and locked before test set evaluation. For trial-level waveform models, participant-level predictions were obtained by aggregating trial-level probabilities within participant using majority vote; participant-level ERP-feature models generated one probability per participant. Test set performance was evaluated at the participant level using AUROC and AUPRC as threshold-independent metrics, and accuracy, sensitivity, specificity, and F1 score at the locked threshold. Ninety-five percent confidence intervals were estimated using 1,000 non-parametric bootstrap replicates resampled at the participant level. Pointwise bootstrap confidence bands for the participant-level receiver-operating-characteristic and precision–recall curves, based on the same 1,000 participant-level resamples, are presented in Supplementary Figures 1 and 2. Additional thresholding and bootstrap implementation details are provided in the Supplementary Methods F.

### Software and hardware

EEG recording and preprocessing was done using Neuroscan QuikCaps and Neuroscan Scan 4.5. Models were implemented in Python 3.12 using PyTorch 2.8.0 with PyTorch Lightning 2.5.2 for the deep learning track and scikit-learn 1.7.1 for the classical track; Optuna 4.4.0 provided Tree-structured Parzen Estimator sampling and persistent SQLite-backed study storage for reproducibility and resumable tuning. Deep learning training used a single CUDA GPU; classical training used multi-core CPU via scikit-learn’s native parallelism.

## Results

### Demographics and grand-average ERP measures

Demographic characteristics and grand average ERP measures stratified by diagnostic group are presented in Table 1. Demographics stratified by train-validation-test splits are presented in Supplementary Table 5. Overall, patients with schizophrenia were younger, had fewer years of education, and had higher smoking rates than healthy controls. ERP measures showed reduced MMN amplitude (−1.35 µV vs. −1.79 µV, p < 0.001) and P3a amplitude (2.43 µV vs. 3.68 µV, p < 0.001) in patients with schizophrenia, consistent with prior literature. Latency differences were modest but statistically significant for both components.

**Table 1.** Demographics and ERP measure grand averages by diagnosis groups.

|  | **Control**<br>**(n=142)** | **Schizophrenia**<br>**(n=258)** | **P** | **Cohen's d** |
| --- | --- | --- | --- | --- |
| Male/Female | 63/79 | 129/129 | 0.28(chi square) |  |
| Age (y/o) | 37.0 ($\pm 11.2$) | 33.0 ($\pm 9.6$) | <0.001 | 0.385 |
| Education (year) | 15.8 ($\pm 2.4$) | 14.0 ($\pm 2.7$) | <0.001 | 0.696 |
| Smoking (PPD) | 0.03 ($\pm 0.15$) | 0.17 ($\pm 0.45$) | <0.001 | -0.377 |
| Onset Age (y/o) | | 24.1 ($\pm 7.1$) | | |
| Duration of illness (year) | | 9.6 ($\pm 9.5$) | | |
| PANSS | | 53.6 ($\pm 15.0$) | | |
| Fz MMN Latency (msec) | 155.2 ($\pm 29.4$) | 148.4 ($\pm 31.6$) | 0.037 | 0.218 |
| Fz MMN Amplitude (uv) | -1.79 ($\pm 0.95$) | -1.35 ($\pm 0.81$) | <0.001 | -0.512 |
| Fz P3a Latency (msec) | 270.7 ($\pm 23.0$) | 264.6 ($\pm 27.6$) | 0.026 | 0.234 |
| Fz P3a Amplitude (uv) | 3.68 ($\pm 1.83$) | 2.43 ($\pm 1.34$) | <0.001 | 0.82 |

### Demographic residualization

On this dataset, demographic covariates explained a median ∼2.4% of subject-mean voltage variance per (channel × time) cell and ∼5.7% across the 18 ERP summary features (maxima 18% at frontal P3a latencies/amplitudes).

### Main Model Metrics: DL models with full residualized ERP waveforms and classical ML models using residualized ERP summary features

Test set threshold-independent metrics (AUROC and AUPRC) of all models are shown in Figure 1. The full model metrics of the deep learning models trained on full residualized EEG waveforms and classical machine learning models trained on 18 ERP summary features are presented in Table 2. In general, deep learning models trained on full epochs demonstrated higher point estimates of classification performance than non-DL machine learning models trained on averaged features. Across the deep learning models, AUROC ranged from 0.797 to 0.857 and AUPRC ranged from 0.854 to 0.906, compared with AUROC values of 0.705 to 0.720 and AUPRC values of 0.813 to 0.831 among the non-DL machine learning models. The autoencoder achieved the highest AUROC among the deep learning models at 0.857 (95% CI 0.767, 0.926), whereas the MLP achieved the highest AUPRC at 0.906 (95% CI 0.838, 0.955). Among the non-DL models, random forest achieved the highest AUROC and AUPRC. Threshold-dependent metrics, including accuracy, sensitivity, specificity, and F1 score, are reported as secondary operating-point summaries given their dependence on the selected threshold. These metrics showed broadly similar patterns favoring deep learning models, although confidence intervals overlapped across models.

**Figure 1.**
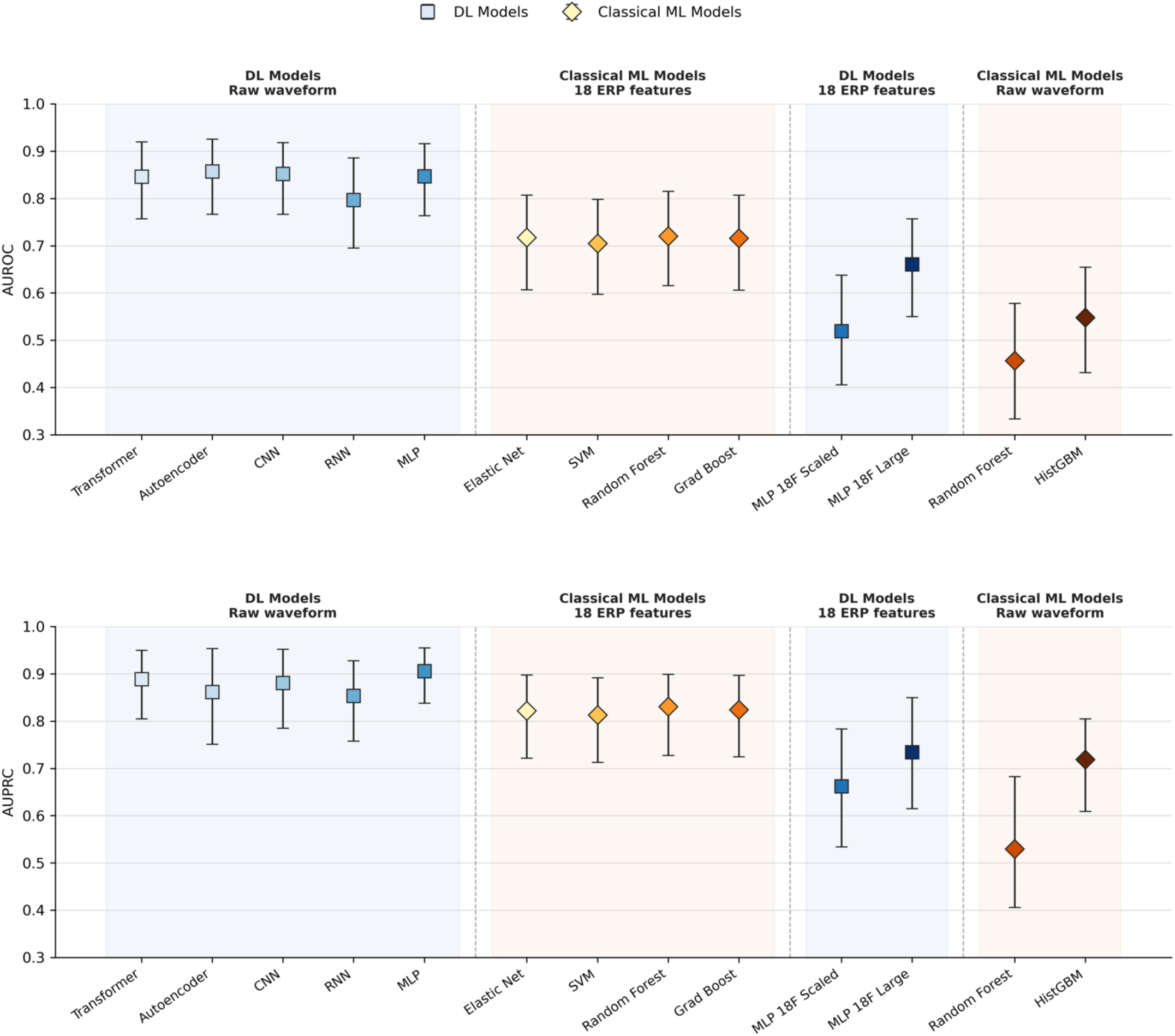
Test-set discrimination performance across waveform and ERP-summary model families. AUROC (top) and AUPRC (bottom) point estimates are shown with 95% participant-level bootstrap confidence intervals. Models are grouped left-to-right as deep learning models trained on full residualized waveforms, classical machine learning models trained on 18 residualized ERP summary features, MLP models trained on the same 18 ERP features, and classical machine learning models trained on full residualized waveforms. Square markers denote deep learning models and diamond markers denote classical machine learning models. Blue and yellow/orange gradients distinguish individual models within each family.

**Table 2.**
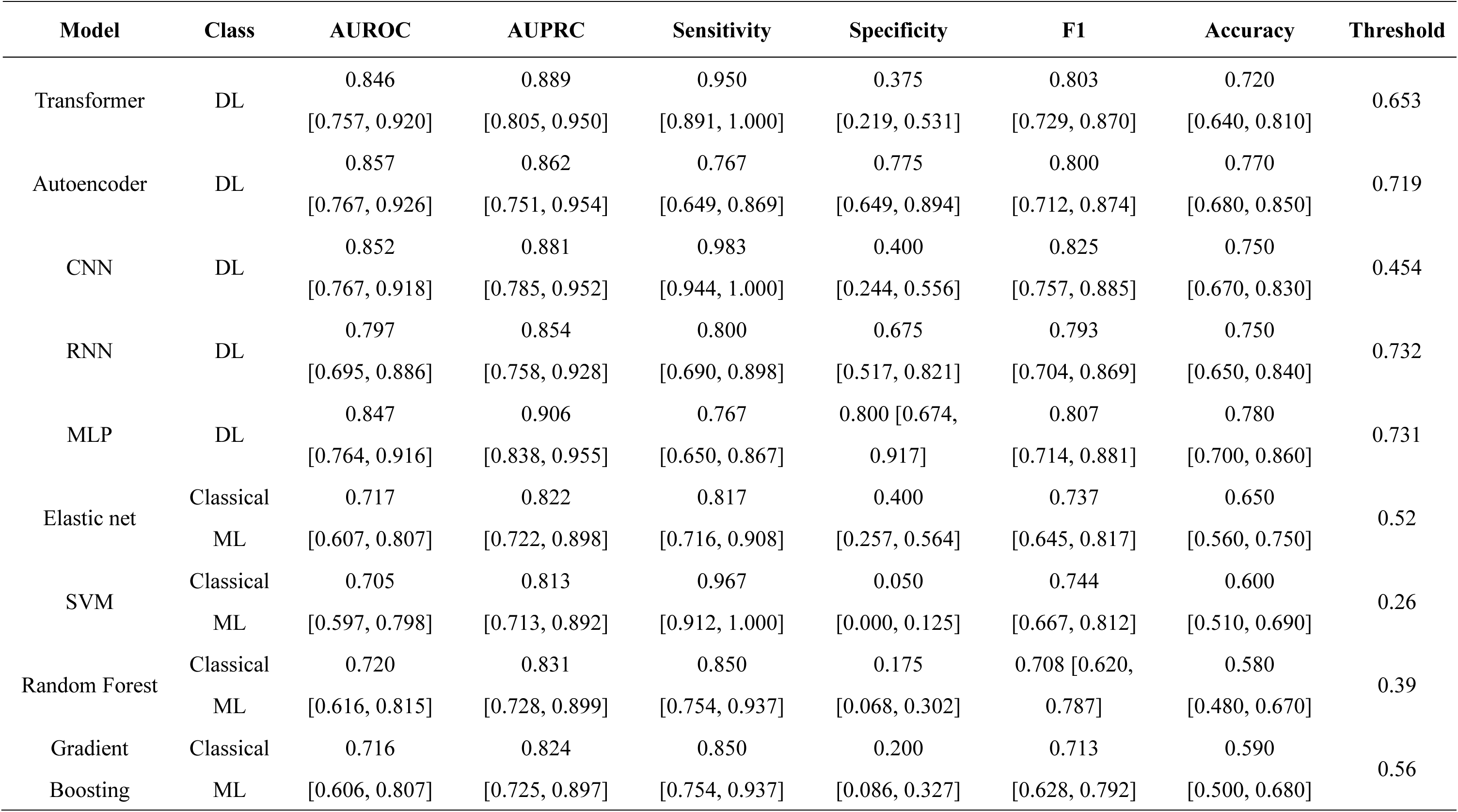
Full model metrics for deep learning models trained on full residualized EEG waveforms and classical machine learning models trained on 18 residualized ERP summary features, reported on the hold-out test set (threshold is obtained by maximum accuracy on the validation set). Values in brackets are 95% confidence intervals based on 1,000 participant-level bootstrap resamples.

### MLP with residualized ERP summary Features

Results from the MLP probe trained on the 18 residualized ERP summary features are shown in Table 3. Overall, the summary-feature MLP models showed lower discrimination than the waveform-based deep learning models. The scaled MLP variant achieved an AUROC of 0.519 (95% CI 0.406, 0.638) and AUPRC of 0.662 (95% CI 0.534, 0.784), whereas the large MLP variant performed better, with an AUROC of 0.661 (95% CI 0.550, 0.757) and AUPRC of 0.734 (95% CI 0.615, 0.850). Despite this improvement, performance remained below that of the waveform-based deep learning models, suggesting that the observed deep learning advantage was not attributable to neural network architecture alone but likely reflected the richer information retained in the full epoch-level waveform inputs.

**Table 3.**
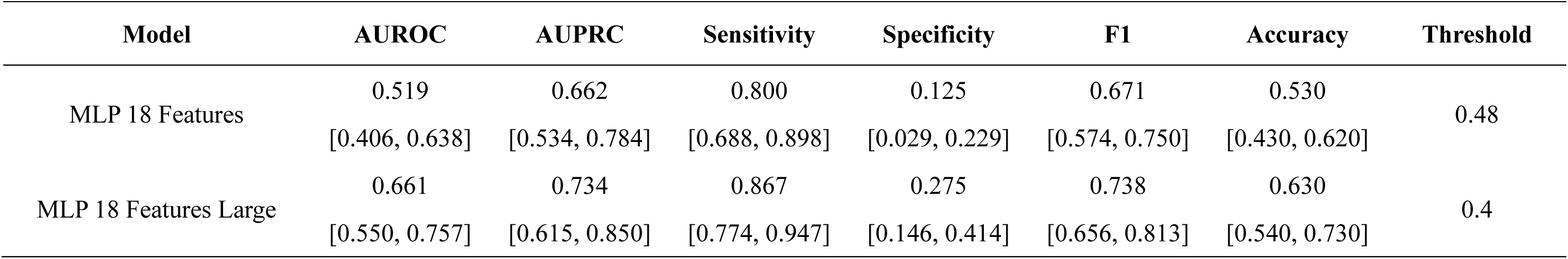
Test-set performance of the scaled and large multi-layer perceptron (MLP) variants trained on 18 residualized ERP summary features. Values in brackets are 95% confidence intervals based on 1,000 participant-level bootstrap resamples.

| Model | AUROC | AUPRC | Sensitivity | Specificity | F1 | Accuracy | Threshold |
| --- | --- | --- | --- | --- | --- | --- | --- |
| MLP 18 Features | 0.519 | 0.662 | 0.800 | 0.125 | 0.671 | 0.530 | 0.48 |
|  | [0.406, 0.638] | [0.534, 0.784] | [0.688, 0.898] | [0.029, 0.229] | [0.574, 0.750] | [0.430, 0.620] |  |
| MLP 18 Features Large | 0.661 | 0.734 | 0.867 | 0.275 | 0.738 | 0.630 | 0.4 |
|  | [0.550, 0.757] | [0.615, 0.850] | [0.774, 0.947] | [0.146, 0.414] | [0.656, 0.813] | [0.540, 0.730] |  |

### Classical ML on full residualized waveforms

Results from the classical machine learning models trained on full residualized waveforms are shown in Table 4. Despite using the same waveform input representation as the deep learning models, the tree-based ensemble models showed limited threshold-independent discrimination. Random forest performed at or below chance-level discrimination, with an AUROC point estimate below 0.5 and a confidence interval spanning chance (AUROC = 0.457, 95% CI 0.334– 0.578; AUPRC = 0.530, 95% CI 0.406–0.683). Histogram-based gradient boosting performed better, with an AUROC of 0.548 (95% CI 0.432–0.655) and AUPRC of 0.719 (95% CI 0.609– 0.805). However, both models remained below the waveform-based deep learning models, and their AUROC confidence intervals included 0.5, indicating that they did not show reliable test-set discrimination in this high-dimensional waveform setting. These results suggest that, among the classical tree-based approaches tested, access to the full epoch-level waveform representation alone was insufficient to reproduce the deep learning advantage. Penalized logistic regression and linear support vector models were also attempted as full-waveform comparators but did not complete fitting or hyperparameter tuning within the available compute budget; detailed feasibility results are provided in the Supplementary Methods.

**Table 4.** Classical ML on full residualized waveforms (n=2; models without convergence issues). Values in brackets are 95% confidence intervals based on 1,000 participant-level bootstrap resamples. Penalized logistic regression and linear SVM did not complete fitting within the available compute budget.

| Model | AUROC | AUPRC | Sensitivity | Specificity | F1 | Accuracy | Threshold |
| --- | --- | --- | --- | --- | --- | --- | --- |
| Random Forest | 0.457 | 0.530 | 0.883 | 0.050 | 0.702 | 0.550 | 0 |
|  | [0.334, 0.578] | [0.406, 0.683] | [0.803, 0.963] | [0.000, 0.128] | [0.620, 0.780] | [0.460, 0.650] |  |
| Hist Gradient Boosting | 0.548 | 0.719 | 0.950 | 0.100 | 0.745 | 0.610 | 0 |
|  | [0.432, 0.655] | [0.609, 0.805] | [0.889, 1.000] | [0.022, 0.209] | [0.662, 0.818] | [0.510, 0.710] |  |

## Discussion

In this study, we evaluated whether auditory oddball ERP waveforms could support individual-level classification of schizophrenia using deep learning models trained on minimally processed trial-level waveform data. Several main findings emerged. First, deep learning models trained on full residualized ERP waveforms achieved higher threshold-independent discrimination than classical machine learning models trained on conventional ERP summary features. Across the deep learning models, AUROC values ranged from 0.797 to 0.857, compared with 0.705 to 0.720 for the classical models trained on 18 ERP summary features. Second, benchmark analyses suggested that this advantage was not attributable to neural-network architecture alone: MLP models trained on the 18 ERP summary features performed below the waveform-based deep learning models. Conversely, access to the full waveform representation alone was also insufficient for the classical tree-based models to reproduce the performance of deep learning models. Together, these findings suggest that the strongest classification performance arose from the combination of rich waveform-level ERP inputs and models capable of learning non-linear structure from high-dimensional temporal data.

Notably, because sensitivity, specificity, and related classification measures depend on the selected decision threshold, we considered AUROC and AUPRC the primary metrics for comparing model discrimination. AUPRC complements AUROC by emphasizing precision and sensitivity for the positive class and should be interpreted relative to the 60% schizophrenia prevalence in the test set, corresponding to a baseline AUPRC of 0.60. The reported threshold-dependent measures reflect a single operating point selected by maximizing participant-level accuracy in the validation set and then applied unchanged to the held-out test set. They should therefore not be interpreted as fixed properties of the models, because alternative thresholds could increase specificity at the cost of sensitivity, or vice versa. Accordingly, differences in specificity at the selected thresholds do not necessarily indicate corresponding differences in overall discrimination, which are more appropriately evaluated using AUROC and AUPRC.

A key strength of the present study is the use of a comparatively large case-control sample relative to much of the prior EEG-based schizophrenia classification literature. Earlier machine learning studies in psychiatry and EEG-based schizophrenia classification have often relied on small cohorts, public datasets, or highly processed feature representations, which can increase the risk of overfitting and inflated performance estimates^6,10,38,39^. By training and evaluating models in 400 participants, including 258 patients with schizophrenia and 142 healthy controls, the present study provides a larger-scale test of whether auditory oddball ERP signals contain information relevant to individual-level case-control classification. The resulting performance estimates were more modest than those reported for some EEG classifiers developed using smaller samples. However, they may provide a more realistic estimate of achievable discrimination given the greater sample heterogeneity and the use of a rigorous participant-level training, validation, and test design. All models used the same participant-level split, and residualization, hyperparameter selection, and decision-threshold locking were performed without access to the held-out test set, thereby reducing the risks of information leakage and overly optimistic performance estimates^10,11,40^. The findings may also be less affected by the small-study effects and selective reporting that can contribute to inflated performance estimates in the published literature. Notably, although our sample of 400 participants was modest for a deep-learning study, each participant completed more than 2,000 trials, with deviant stimuli occurring with a 10% probability. This design yielded a large number of deviant-trial waveforms per participant, although the realized number of deviants and the number retained after artifact rejection varied across participants. These repeated measurements provided substantial within-participant waveform information for representation learning and may have supported the observed discrimination performance.

A central motivation for this study was the distinction between group-level ERP abnormalities and individual-level prediction. MMN and P3a deficits are among the most consistently reported electrophysiological abnormalities in schizophrenia and are commonly interpreted as markers of impaired sensory prediction, deviance detection, and attentional orienting^16^. However, robust group differences do not necessarily imply clinically useful single-subject classification^9^. Conventional ERP analyses typically summarize averaged waveforms using peak amplitudes, latencies, or mean area measures. Although these features are interpretable and have strong neurophysiological grounding, they may discard temporal morphology, distributed spatial structure, and trial-level variability that could be informative for participant-level prediction^27^. Our findings support this possibility: models trained on full trial-level waveforms outperformed models trained on conventional ERP summary features, suggesting that schizophrenia-related electrophysiological information may not be fully captured by canonical MMN and P3a summary measures alone.

The use of auditory oddball ERPs also has translational relevance beyond classification accuracy alone. Compared with EEG representations that are not explicitly anchored to a specific stimulus or cognitive process, stimulus-locked ERPs are experimentally controlled and tied to defined neurocognitive operations^16^. This makes them attractive not only as inputs for classification models, but also as experimentally tractable biomarkers of disease-relevant brain function. In particular, MMN and P3a can be elicited under passive listening conditions, reducing dependence on active task performance and making the paradigm more feasible for patients with cognitive impairment, reduced motivation, or limited task engagement^16^. ERPs are also relatively low-cost, temporally precise, and more scalable than neuroimaging modalities such as MRI, which remain constrained by cost, accessibility, and limited temporal resolution^6,13^. These properties support their potential role in clinical decision-support pipelines, especially if combined with interpretable modeling approaches and external validation.

Although some variation was observed, discrimination performance was broadly similar across the waveform-based deep learning models, despite their differing inductive biases regarding local temporal morphology, sequential dependence, and long-range interactions^41,42^. This finding suggests that, in a moderately large dataset with high-dimensional ERP inputs, substantial discriminative information may be present in distributed waveform patterns that do not require strong architecture-specific inductive biases to be useful. Future work could examine whether ensemble approaches combining these architectures improve calibration, sensitivity, specificity, or robustness across datasets.

Several limitations should be acknowledged. First, the study was conducted at a single site using a single acquisition system and auditory oddball paradigm, limiting generalizability across populations, recording systems, and preprocessing pipelines. External validation in independent cohorts will therefore be essential. Second, the classification task was limited to patients with schizophrenia versus healthy controls. This design tests case-control discriminability but does not establish diagnostic specificity relative to other psychiatric or neurodevelopmental conditions. Future studies should include differential-diagnosis cohorts, including bipolar disorder, major depressive disorder, and clinical high-risk populations. Third, although we benchmarked deep learning models trained on full waveform inputs against classical machine learning and deep learning models trained on conventional ERP summary features, this comparison was not exhaustive. In particular, we did not evaluate every potentially useful handcrafted feature set or dimensionality-reduction strategy. Nevertheless, our findings suggest that deep learning models can learn useful representations directly from minimally processed ERP waveforms, reducing reliance on manual feature design, which remains a major bottleneck in many classical machine learning approaches. Fourth, illness heterogeneity, medication exposure, illness duration, symptom dimensions, smoking burden, and cognitive functioning may all influence ERP morphology and classification performance. Future work should examine whether model performance varies across clinical subgroups or illness stages. Fifth, interpretability remains a major challenge^1^. Although the present findings support the potential utility of trial-level ERP waveforms for case-control classification, clinical adoption will require transparent links between model outputs and neurophysiological processes. Explainable-AI approaches, such as saliency maps, occlusion analyses, attention visualization, or temporal-channel attribution, may help identify which portions of the MMN or P3a time windows contribute most strongly to classification decisions^41^.

In conclusion, this study suggests that auditory oddball ERP waveforms contain participant-level information relevant to schizophrenia classification that is not fully captured by conventional averaged ERP summary features. Deep learning models applied to full trial-level waveforms showed higher discrimination than conventional feature-based approaches in a comparatively large case-control sample, supporting the value of preserving ERP temporal and spatial structure for predictive modeling. More broadly, auditory oddball ERPs may serve as candidate classification inputs and experimentally tractable biomarkers of disease-relevant neural information processing, either alone or in combination with complementary modalities such as structural MRI^6^ or polygenic risk scores^12^. With external validation, improved interpretability, and evaluation in differential-diagnosis cohorts, ERP-based deep learning approaches may contribute to scalable and biologically informed tools for precision psychiatry.

## Data Availability

The individual-level EEG and clinical data analyzed in this study cannot be made publicly available because the participants’ informed consent and the governing institutional review board approval do not authorize public data sharing. Access to these data is therefore restricted to the approved study team. Aggregate results supporting the findings are reported in the article and its supplementary materials.

## Acknowledgements

This work was supported in part by the Taiwan National Science and Technology Council (NSTC) under grants NSTC 113-2314-B-002-185-MY2 and NSTC 111-2314-B-002-102-MY2.

