## Supplementary materials for "Deep Learning for Individual-Level Classification of Schizophrenia Versus Healthy Controls from Trial-Level Auditory Oddball ERP Waveforms"

**Supplementary Methods A.** Implementation details of demographic residualization

**Supplementary Methods B.** Implementation details of deep learning models on full residualized waveforms

**Supplementary Methods C.** Implementation details of classical machine learning models on 18 residualized ERP summary features

**Supplementary Methods D.** Implementation details of multi-layer perceptron probe on 18 residualized ERP summary features

**Supplementary Methods E.** Implementation details classical machine learning models on full residualized waveforms

**Supplementary Methods F.** Implementation details of threshold selection, Evaluation metrics, and Uncertainty quantification

**Supplementary Figure 1.** Participant-level ROC curves with pointwise 95% confidence bands based on 1,000 bootstrap resamples of the held-out test participants.

**Supplementary Figure 2.** Participant-level PR curves with pointwise 95% confidence bands based on 1,000 bootstrap resamples of the held-out test participants.

**Supplementary Table 1.** Hyperparameter search spaces and selected configurations for deep learning models trained on full residualized waveforms.

**Supplementary Table 2.** Hyperparameter grids and selected configurations for classical machine learning models trained on 18 residualized ERP summary features.

**Supplementary Table 3.** Hyperparameter search spaces and selected configurations for multi-layer perceptrons trained on 18 residualized ERP summary features.

**Supplementary Table 4.** Hyperparameter grids and selected configurations for classical machine learning models trained on full residualized waveforms.

**Supplementary Table 5.** Demographics of the 400 participants by train-validation-test data split.

**Supplementary Methods A.** Implementation details of demographic residualization

To reduce the risk that classifiers would exploit demographic differences between patients with schizophrenia and healthy controls rather than illness-specific neurophysiology, we residualized the EEG features used by every downstream model against four demographic covariates before training: age in years, sex coded as binary 0/1 after recoding from the collected 1/2 variable, years of education, and packs of cigarettes per day. One participant with schizophrenia had a missing education value; this value was imputed using the median education of participants with schizophrenia in the training split only. The imputed value was used only for residualization and was not persisted back to the source data file.

Residualization was implemented as a participant-level procedure. For each output dimension—either each channel × time cell for full waveforms used by deep learning models or each summary-feature column for the ERP summary-feature set—we fit an ordinary least-squares regression on the 250 training participants' subject-mean response against the four covariates, including an intercept. The resulting regression coefficients were then held fixed and applied to every trial of every participant in the training, validation, and test splits by broadcasting the per-subject predicted demographic component and subtracting it from the original signal.

This procedure ensured that validation and test data were never used to estimate residualization coefficients, thereby preventing information leakage. It also ensured that every trial from a given participant received the same demographic correction, rather than allowing trial-specific noise to be modeled as demographic variation. Participant-level coefficients, covariate means, per-cell $R^2$

values, and pre-/post-residualization covariate-correlation diagnostics were archived with the model outputs.

**Supplementary Methods B.** Implementation details of deep learning models on full residualized waveforms

Five deep learning architectures were trained on trial-level residualized EEG waveforms. The Transformer used positional encoding followed by multi-head self-attention. The autoencoder used a convolutional encoder-decoder with a joint reconstruction and classification objective, weighted by a scalar alpha. The convolutional neural network (CNN) used stacked one-dimensional temporal convolutions. The recurrent neural network (RNN) search included LSTM, GRU, and vanilla RNN units, with optional bidirectionality and attention. The multi-layer perceptron (MLP) was applied to the flattened 21,636-feature waveform input.

All five models used a log-softmax output layer with negative-log-likelihood loss and the Adam optimizer with default beta parameters. Linear layers used Xavier-uniform initialization with zeroed biases. Gradients were clipped to an L2 norm of 0.5, and input values were clamped to the range -10 to 10 for numerical stability. Batch size was 64. The RNN used gradient accumulation over two batches; the other models updated parameters after every batch.

Hyperparameters were selected using Optuna with the default Tree-structured Parzen Estimator sampler. Each architecture was evaluated in 25 trials, with a maximum of 30 epochs per trial and early-stopping patience of three validation epochs. Dropout was sampled uniformly from 0.1 to 0.4, and the learning rate was sampled log-uniformly from 1 x 10^-5 to 5 x 10^-5 for every architecture. Neural-network initialization and training used random seed 36; an explicit random seed was not assigned to the Optuna sampler. The selection objective was validation participant-level AUROC calculated using majority-vote aggregation of trial-level predictions. The checkpoint from the epoch with the highest validation participant-level AUROC in the best trial was retained.

The selected model was not retrained on the combined training and validation sets. Because the validation set was used for both hyperparameter selection and decision-threshold selection, retraining would have produced a new fitted model without an independent dataset available for threshold locking. The train-only selected checkpoint was therefore retained, and its validation-locked threshold was applied unchanged to the held-out test set.

The tuned parameters controlled model capacity, temporal processing, and regularization. For the Transformer, the number of attention heads determined how many parallel representation subspaces were used for self-attention; the feed-forward hidden dimension and number of encoder layers controlled the width and depth of the encoder; and the classification-head hidden dimension controlled the capacity of the final decision layers. For the autoencoder, the latent dimension determined the size of the compressed representation. Its joint loss was defined as alpha times the reconstruction mean-squared error plus (1 - alpha) times the classification loss, so larger alpha values placed more weight on waveform reconstruction. Convolutional kernel size determined the temporal receptive field, stride determined temporal downsampling, and filter count determined the number of learned feature maps. For the CNN, the number of layers controlled network depth, filter count controlled channel capacity, kernel size controlled temporal receptive field, pooling size controlled downsampling, and the fully connected hidden dimension controlled the capacity of the classification head. For the RNN, recurrent-unit type specified the state-update mechanism; hidden dimension and layer count controlled recurrent capacity and depth; bidirectionality permitted information to be processed in both temporal directions; attention enabled weighted aggregation across time; and the fully connected hidden dimension controlled the classification head. For the waveform MLP, layer count and width controlled network depth and capacity, whereas batch normalization standardized intermediate activations. Across all neural-network models, dropout specified the proportion of hidden activations randomly set to zero during training, and the learning rate controlled the size of optimizer updates.

**Supplementary Methods C.** Implementation details of classical machine learning models on 18 residualized ERP summary features

Four classical machine learning classifiers were trained using the 250 training participants' 18 residualized ERP summary features: elastic net penalized logistic regression, support vector classification, random forest, and gradient boosting. Logistic regression used an elastic net penalty with the SAGA solver. Support vector classification included linear and radial-basis function kernels with probability estimates enabled. Feature standardization was performed within the cross-validation pipeline for logistic regression and support vector classification, so scaling parameters were estimated using only the applicable training folds. Tree-based models used the residualized feature values without additional scaling.

Hyperparameters were selected by exhaustive grid search using five-fold stratified cross-validation within the training split and AUROC as the scoring metric. Cross-validation shuffling and model initialization used random seed 36. The validation set was not used during grid search and was reserved for decision-threshold selection after the best configuration had been fitted to all 250 training participants. The grids contained 25 configurations for elastic net logistic regression, 16 for support vector classification, 270 for random forest, and 3,888 for gradient boosting. Class-weight tuning was not performed; all models used the default uniform class weighting, consistent with the unweighted loss functions used for the deep learning models.

The tuned parameters controlled regularization and model complexity. For elastic net logistic regression, C was the inverse regularization strength, so larger values imposed weaker overall regularization, while the L1 ratio controlled the balance between sparse L1 and ridge-like L2 penalties. For the support vector classifier, kernel type determined whether the decision boundary was linear or nonlinear; C controlled the penalty for classification errors; and, for the radial-basis function kernel, gamma controlled how locally each training observation influenced the decision boundary. For random forest, the number of trees controlled ensemble size, maximum depth limited individual-tree complexity, minimum split and leaf sizes regularized tree growth, and the maximum-feature setting controlled the number of candidate features considered at each split. For gradient boosting, the number of estimators controlled the number of sequential boosting stages, the learning rate scaled the contribution of each stage, maximum depth and minimum split or leaf sizes controlled the complexity of the component trees, the subsample fraction introduced row subsampling, and the maximum-feature setting introduced feature subsampling.

**Supplementary Methods D.** Implementation details of multi-layer perceptron probe on 18 residualized ERP summary features

To isolate the contribution of model architecture from input richness, we trained PyTorch Lightning multi-layer perceptrons on the 18 participant-level ERP summary features. The models used the same core building blocks as the waveform MLP: stacked linear, batch-normalization, rectified-linear-unit, and dropout layers; Xavier-uniform initialization; a log-softmax output layer; negative-log-likelihood loss; and the Adam optimizer. The 18-feature inputs were standardized using means and standard deviations estimated from the training split. Input clamping was not applied because the summary features were not on the waveform voltage scale.

Two search-space variants were evaluated. The scaled variant restricted hidden-layer widths to 16, 32, 64, or 128 units across one to three layers. This search space was designed for the relatively small 250-participant, 18-feature training set and reduced the degree of overparameterization. The large variant allowed widths from 128 to 1024 units across two to four layers, matching the search space of the waveform MLP. This variant tested whether the larger waveform-oriented architecture impaired performance when applied to low-dimensional summary features.

Each variant was tuned with 25 Optuna trials using the default Tree-structured Parzen Estimator sampler. Dropout was sampled uniformly from 0.1 to 0.4 and the learning rate log-uniformly from 1 x 10^-5 to 5 x 10^-5. Trials were trained for up to 50 epochs with early-stopping patience of 10 validation epochs and batch size 64. Neural-network initialization and training used random seed 36; an explicit random seed was not assigned to the Optuna sampler. The selection objective was validation participant-level AUROC. The checkpoint from the epoch with the highest validation AUROC in the best trial was retained and evaluated on the held-out test set without retraining on the combined training and validation sets.

For these MLPs, the number of hidden layers and the width of each layer controlled network depth and representational capacity. Batch normalization standardized intermediate activations and could stabilize optimization, dropout regularized the network by randomly setting a proportion of hidden activations to zero during training, and the learning rate controlled the size of Adam optimizer updates.

**Supplementary Methods E.** Implementation details of classical machine learning models on full residualized waveforms

For direct comparison with the deep learning models using the same input representation, two tree-based classical ensemble models were trained at the trial level on flattened 21,636-dimensional residualized waveforms: random forest and histogram-based gradient boosting. Histogram-based gradient boosting used quantile pre-binning into 255 bins followed by histogram-based split finding, making it computationally tractable for approximately 75,000 training trials with 21,636 features. Conventional gradient boosting was not attempted at this scale because of its substantially greater computational demands.

Hyperparameters were selected using five-fold stratified group cross-validation within the training split. Folds were grouped by participant so that trials from the same participant could not appear in both the training and held-out portions of a fold. Model selection used a custom scoring function that calculated participant-level majority-vote AUROC on each held-out fold, matching the aggregation strategy used for the waveform deep learning models. Cross-validation shuffling and model initialization used random seed 36. Each tree-based model had 27 candidate configurations, providing a search breadth similar to the 25-trial Optuna budget used for each deep learning model. After selection, the best configuration was fitted to the complete training split; the separate validation split was then used to lock the decision threshold.

For random forest, the number of trees controlled ensemble size, maximum depth limited the complexity of each tree, minimum leaf size prevented splits that produced very small terminal nodes, and square-root feature sampling limited the number of candidate features considered at each split. For histogram-based gradient boosting, the learning rate scaled the contribution of each sequential boosting stage, maximum iterations limited the number of stages, and maximum depth constrained the complexity of the component trees; an unlimited depth allowed growth to continue subject to the estimator's other stopping rules.

Penalized logistic regression and linear support vector classification were also attempted as full-waveform comparators but were not included in the final quantitative comparison because they were not computationally feasible within the available compute budget. Elastic net logistic regression with the SAGA solver did not complete its 20-fit hyperparameter search after 9 hours 25 minutes of CPU time and produced convergence warnings. Linear support vector classification with sigmoid probability calibration did not complete its first fit after 4 hours 26 minutes. These failures are consistent with the scalability limitations of gradient-based and coordinate-descent linear solvers when applied to high-dimensional matrices with strongly correlated features. Future work could evaluate dimensionality reduction or stochastic-gradient solvers for linear full-waveform comparisons.

**Supplementary Methods F.** Implementation details of threshold selection, Evaluation metrics, and Uncertainty quantification

*Threshold selection*

All models operate on probabilistic output. For each model we selected a single decision threshold after training by maximizing participant-level accuracy on the validation split, with a deterministic tie-break (lowest threshold among tied values) and the resulting threshold was locked and applied unchanged to the test split. Candidate thresholds were defined according to prediction granularity: participant-level ERP models used a fixed 0.01-spaced grid, whereas trial-level waveform models swept empirical trial-probability breakpoints, capped for computational tractability. For trial-level models (the five waveform deep models and the trial-level classical waveform models), participant-level predictions were obtained by majority-vote aggregation over each participant's trials: the proportion of trials for which the positive-class probability exceeded the threshold was compared to 0.5, and the participant was assigned the majority label. For participant-level ERP-feature models, each participant contributed one feature vector and therefore one positive-class probability, which was thresholded directly.

*Evaluation metrics*

For each model, performance was evaluated on the held-out test set. Threshold-free discrimination was assessed using the area under the receiver-operating-characteristic curve (AUROC) and the area under the precision-recall curve (AUPRC), both computed at the patient level. For trial-level models, participant-level predictions were obtained using the majority-vote aggregation procedure described above, whereas participant-level ERP-feature models used the single predicted probability generated for each participant. We also report operating-point metrics at the locked classification threshold, including accuracy, sensitivity, specificity, precision, negative predictive value, and F1 score.

*Uncertainty quantification*

95% confidence intervals for every reported metric were computed by non-parametric bootstrap at the participant level, not at the trial level: 1,000 bootstrap replicates of the 100 test participants were drawn with replacement. For trial-level models, each replicate reconstructed the corresponding trial-level dataset by concatenating all trials of the sampled participants; participants drawn multiple times contributed multiple copies of their full trial set, treated as distinct participants for aggregation. For participant-level ERP-feature models, the same resampling was performed over one probability per participant. In each replicate, AUROC and AUPRC were recomputed across the full threshold sweep, whereas threshold-dependent metrics were recomputed using the locked validation-selected threshold. The 2.5th and 97.5th percentiles of the resulting 1,000-sample bootstrap distribution formed the confidence interval. The same participant-level bootstrap procedure was used to construct pointwise 95% confidence bands for the participant-level ROC and precision–recall curves. Within each replicate, repeated false-positive-rate or recall coordinates were resolved by retaining the maximum corresponding true-positive rate or precision, respectively, and the curves were linearly interpolated onto common 101-point grids. The 2.5th and 97.5th percentiles at each grid value formed the pointwise confidence bands. Model parameters, residualization coefficients, and hyperparameters were held fixed; model fitting and selection were not repeated during bootstrap resampling. Resampling at the participant level preserves the strong within-subject correlation structure of EEG and avoids the well-documented under-estimation of uncertainty that trial-level resampling produces when correlated observations are treated as independent. The RNG seed was fixed (42) so confidence intervals are reproducible.

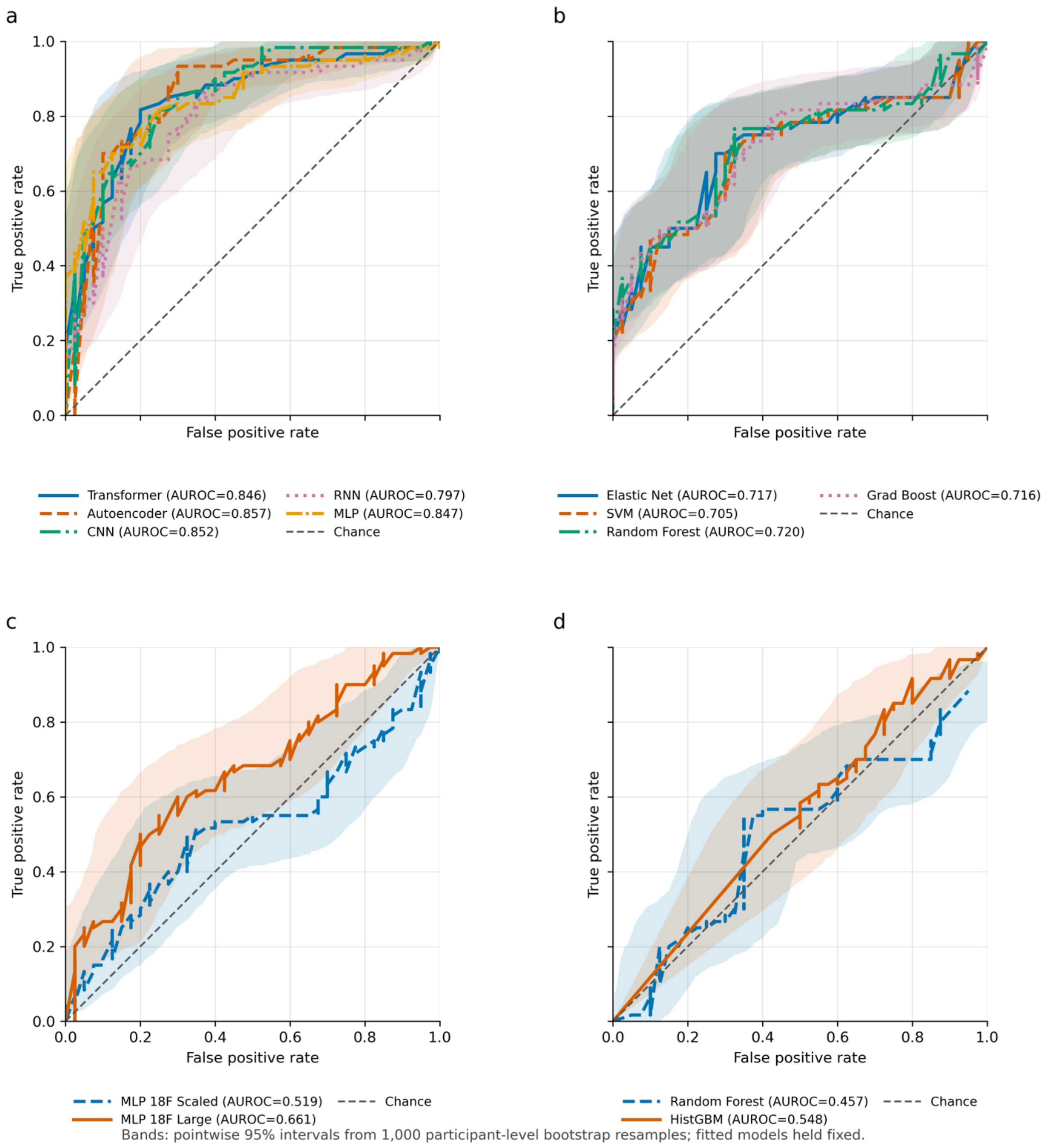


**Supplementary Figure 1.** Participant-level ROC curves with pointwise 95% confidence bands based on 1,000 bootstrap resamples of the held-out test participants. Receiver operating characteristic (ROC) curves are shown for (a) five deep learning models trained on full residualized waveforms, (b) four classical machine learning models trained on 18 residualized ERP summary features, (c) two deep learning MLP variants trained on the same 18 ERP features, and (d) two classical machine learning models trained on full residualized waveforms. Line color and line style jointly identify each model; legend values are participant-level AUROC estimates. Shaded regions represent pointwise 95% confidence bands derived from 1,000 participant-level bootstrap resamples of the held-out test set, as described in Supplementary Methods F. The dashed diagonal denotes chance discrimination.

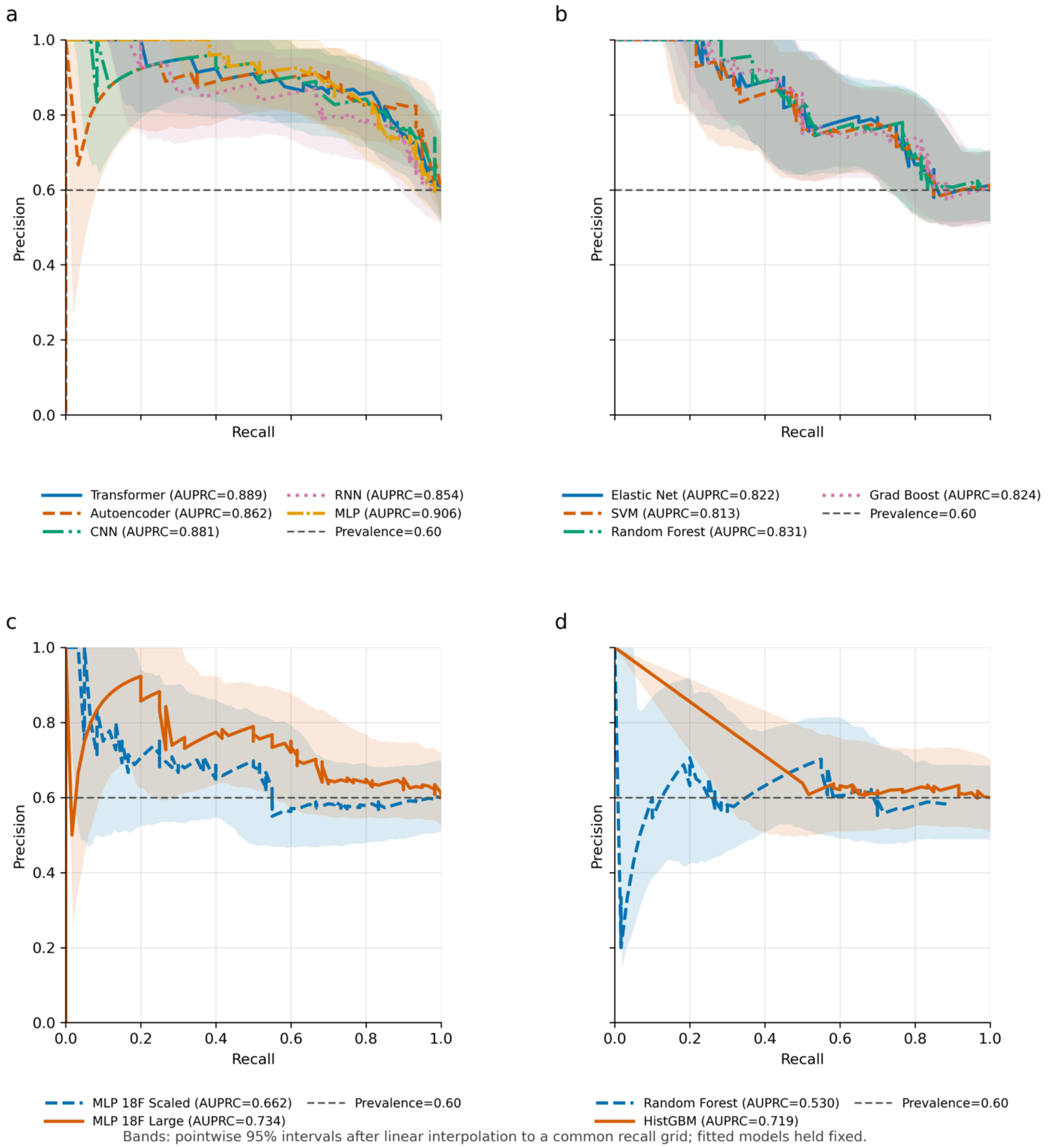


**Supplementary Figure 2.** Participant-level PR curves with pointwise 95% confidence bands based on 1,000 bootstrap resamples of the held-out test participants. Precision-recall curves are shown for (a) five deep learning models trained on full residualized waveforms, (b) four classical machine learning models trained on 18 residualized ERP summary features, (c) two deep learning MLP variants trained on the same 18 ERP features, and (d) two classical machine learning models trained on full residualized waveforms. Line color and line style jointly identify each model; legend values are participant-level AUPRC estimates. Empirical curves linearly connect the majority-vote threshold-sweep coordinates. Shaded regions represent pointwise 95% confidence bands derived from 1,000 participant-level bootstrap resamples of the held-out test set, as described in Supplementary Methods F. The dashed horizontal line denotes the schizophrenia prevalence in the test set (0.60).

**Supplementary Table 1.** Hyperparameter search spaces and selected configurations for deep learning models trained on full residualized waveforms.

| Model | Search space | Selected configuration |
|---|---|---|
| Transformer | Attention heads: {3, 6, 12}; feed-forward hidden dimension: 128-512 in steps of 64; encoder layers: 2-4; classification-head hidden dimension: 64-256 in steps of 32; dropout and learning rate as described above. | 12 attention heads; feed-forward hidden dimension 192; 3 encoder layers; classification-head hidden dimension 256; dropout 0.255467; learning rate 1.149687 x 10^-5. |
| Autoencoder | Latent dimension: 64-256 in steps of 32; alpha: 0.3-0.7, sampled uniformly; first and second convolutional kernel sizes: {5, 10, 15, 20}; first and second strides: {1, 2, 3}; first-layer filters: {8, 16, 32}; second-layer filters: {16, 32, 64}; dropout and learning rate as described above. | Latent dimension 192; alpha 0.307274; kernel sizes 5 and 20; strides 2 and 2; filter counts 8 and 64; dropout 0.214827; learning rate 1.031874 x 10^-5. |
| CNN | Convolutional layers: 3-5; fully connected hidden dimension: 128-512 in steps of 64; for every included layer, base filters: {8, 16, 32}, kernel size: {3, 5, 7}, and pooling size: {1, 2, 3}. The number of filters at layer i was the base filter count multiplied by 2^i; dropout and learning rate as described above. | 3 convolutional layers; fully connected hidden dimension 384; base filters 16, 16, and 16, corresponding to actual filter counts 16, 32, and 64; kernel sizes 7, 5, and 7; pooling sizes 3, 1, and 2; dropout 0.243737; learning rate 1.364998 x 10^-5. |
| RNN | Recurrent unit: {LSTM, GRU, vanilla RNN}; hidden dimension: 64-256 in steps of 32; recurrent layers: 1-4; bidirectionality: {enabled, disabled}; fully connected hidden dimension: 128-512 in steps of 64; attention: {enabled, disabled}; dropout and learning rate as described above. | LSTM; hidden dimension 160; 3 recurrent layers; bidirectionality enabled; fully connected hidden dimension 320; attention enabled; dropout 0.225447; learning rate 2.004942 x 10^-5. |
| Waveform MLP | Hidden layers: 2-4; width of each included layer: 128-1024 in steps of 128; batch normalization: {enabled, disabled}; dropout and learning rate as described above. | 2 hidden layers with 256 and 768 units; batch normalization disabled; dropout 0.203534; learning rate 3.000304 x 10^-5. |

**Supplementary Table 2.** Hyperparameter grids and selected configurations for classical machine learning models trained on 18 residualized ERP summary features.

| Model | Hyperparameter grid | Selected configuration |
|---|---|---|
| Elastic net logistic regression | Inverse regularization strength C: {0.01, 0.1, 1, 10, 100}; L1 ratio: {0.1, 0.3, 0.5, 0.7, 0.9}; elastic net penalty; SAGA solver; maximum 5,000 iterations. Total: 25 configurations. | C = 0.1; L1 ratio = 0.1. |
| Support vector classifier | Linear kernel: C in {0.01, 0.1, 1, 10}. Radial-basis function kernel: the same four C values crossed with gamma in {scale, 0.01, 0.1}. Probability estimates enabled. Total: 16 configurations. | Linear kernel; C = 0.1. |
| Random forest | Trees: {50, 100, 200}; maximum depth: {2, 3, 5, 7, 10}; minimum samples per split: {5, 10, 20}; minimum samples per leaf: {5, 10, 15}; maximum features: {square root, log2}. Total: 270 configurations. | 200 trees; maximum depth 2; minimum samples per split 20; minimum samples per leaf 5; square-root feature sampling. |
| Gradient boosting | Estimators: {50, 100, 150}; learning rate: {0.01, 0.05, 0.1, 0.2}; maximum depth: {2, 3, 4, 5}; minimum samples per split: {10, 20, 30}; minimum samples per leaf: {5, 10, 15}; subsample fraction: {0.7, 0.8, 0.9}; maximum features: {square root, log2, 0.5}. Total: 3,888 configurations. | 50 estimators; learning rate 0.01; maximum depth 2; minimum samples per split 30; minimum samples per leaf 10; subsample fraction 0.8; square-root feature sampling. |

**Supplementary Table 3.** Hyperparameter search spaces and selected configurations for multi-layer perceptrons trained on 18 residualized ERP summary features.

| Model | Search space | Selected configuration |
|---|---|---|
| Scaled 18-feature MLP | Hidden layers: 1-3; width of each included layer: {16, 32, 64, 128}; batch normalization: {enabled, disabled}; dropout: 0.1-0.4, sampled uniformly; learning rate: 1 x 10^-5 to 5 x 10^-5, sampled log-uniformly. | 2 hidden layers with 16 and 32 units; batch normalization enabled; dropout 0.210189; learning rate 2.846397 x 10^-5. |
| Large 18-feature MLP | Hidden layers: 2-4; width of each included layer: 128-1024 in steps of 128; batch normalization: {enabled, disabled}; dropout: 0.1-0.4, sampled uniformly; learning rate: 1 x 10^-5 to 5 x 10^-5, sampled log-uniformly. | 3 hidden layers with 384, 640, and 768 units; batch normalization enabled; dropout 0.350569; learning rate 3.655282 x 10^-5. |

**Supplementary Table 4.** Hyperparameter grids and selected configurations for classical machine learning models trained on full residualized waveforms.

| Model | Search space | Selected configuration |
|---|---|---|
| Random forest | Trees: {50, 100, 200}; maximum depth: {5, 10, unlimited}; minimum samples per leaf: {5, 10, 20}; maximum features: {square root}. Total: 27 configurations. | 200 trees; maximum depth 5; minimum samples per leaf 10; square-root feature sampling. |
| Histogram-based gradient boosting | Learning rate: {0.05, 0.1, 0.2}; maximum iterations: {100, 200, 300}; maximum depth: {unlimited, 5, 10}. Total: 27 configurations. | Learning rate 0.2; 300 iterations; unlimited depth. |

**Supplementary Table 5.** Demographics of the 400 participants by train-validation-test data spl

| Split | Total # | # Controls | # Patients with Schizophrenia | Age (yr, mean ± SD) | Female n (%) | Male n (%) | Education (yr, mean ± SD) | PPD (packs/day, mean ± SD) |
|---|---|---|---|---|---|---|---|---|
| Overall | 400 | 142 (35.5%) | 258 (64.5%) | 34.4 ± 10.3 | 208 (52.0%) | 192 (48.0%) | 14.6 ± 2.7 | 0.1 ± 0.4 |
| Train | 250 | 84 (33.6%) | 166 (66.4%) | 34.4 ± 10.2 | 133 (53.2%) | 117 (46.8%) | 14.8 ± 2.5 | 0.1 ± 0.3 |
| Validation | 50 | 18 (36.0%) | 32 (64.0%) | 35.1 ± 10.1 | 26 (52.0%) | 24 (48.0%) | 13.9 ± 2.4 | 0.2 ± 0.4 |
| Test | 100 | 40 (40.0%) | 60 (60.0%) | 34.3 ± 10.9 | 49 (49.0%) | 51 (51.0%) | 14.4 ± 3.3 | 0.1 ± 0.5 |
